# A single-session randomised crossover fNIRS study comparing three upper-limb mirror therapy task paradigms in healthy adults: a study protocol

**DOI:** 10.64898/2026.08.28.26361691

**Authors:** Tangzhu Yang, Siyuan Wei, Yuezhu Wang, Dingqun Bai

**Affiliations:** Department of Rehabilitation Medicine, Wuchang Hospital Affiliated to Wuhan University of Science and Technology, Wuhan, China; Key Laboratory of Physical Medicine and Precision Rehabilitation of Chongqing Municipal Health Commission, Department of Rehabilitation Medicine, The First Affiliated Hospital of Chongqing Medical University, Chongqing, China

**Keywords:** mirror therapy, mirror visual feedback, functional near-infrared spectroscopy, upper limb, healthy people, task paradigm, crossover design

## Abstract

**Background:** Mirror therapy (MT)—specifically paradigms using mirror visual feedback (MVF)—is widely used in neurorehabilitation; however, mechanistic implementations vary substantially in movement content, rhythmicity and attentional demands. This protocol describes an acute mechanistic, within-participant fNIRS screening study designed to compare three prespecified upper-limb mirror-therapy task paradigms and to quantify associated subjective experience after each condition in healthy adults during a single visit.

**Methods and analysis:** This is a single-centre, within-participant, randomised crossover study conducted at Wuhan Wuchang Hospital (Wuhan, China). Healthy adults aged 18–35 years will complete three task conditions once each in a counterbalanced order using a 3×3 Latin-square scheme: UMT1 (task-oriented rhythmic functional movement), UMT2 (open-ended free movement with auditory control), and UMT3 (non-functional rhythmic movement). fNIRS will be acquired using the NirSmart-6000A system during a standardised block design. The primary outcome is ROI-level HbO activation quantified as GLM-derived β estimates within the prespecified primary ROIs (bilateral SM1/M1 and bilateral PMC). Secondary outcomes include ROI-level windowed ΔHbO (5–20 s post-onset relative to the immediately preceding rest; descriptive only), ROI-level ΔHbR, and post-condition subjective ratings (illusion, immersion, confusion and fatigue; 1–7 Likert). Condition effects will be analysed using linear mixed-effects models with fixed effects for condition and period and prespecified multiplicity-adjusted pairwise contrasts.

**Ethics and dissemination:** Ethics approval was obtained from the Ethics Committee of Wuchang Hospital Affiliated to Wuhan University of Science and Technology (Approval No.: 2025-112-01; approved on 2025-08-21). The study is expected to be minimal risk. Findings will be disseminated through publication of this protocol manuscript and subsequent results manuscripts and conference presentations.

**Trial registration number:** Chinese Clinical Trial Registry (ChiCTR2600116634). This study is conducted as a prespecified mechanistic sub-study under the overarching registered project.

**Strengths and limitations of this study:**

- Within-participant randomised crossover design reduces between-participant variability for mechanistic comparisons.
- Prespecified task timing, ROIs, contrasts and multiplicity control reduce analytic flexibility.
- Latin-square counterbalancing and modelling of period effects mitigate systematic sequence bias.
- Healthy-participant, single-session design limits direct generalisability to clinical stroke populations and long-term treatment effects.
- fNIRS remains sensitive to motion and scalp–optode coupling; residual artefacts may persist despite prespecified QC and preprocessing.

## Introduction

Mirror therapy (MT)—specifically paradigms using mirror visual feedback (MVF)—is widely used in neurorehabilitation and has been associated with neurophysiological modulation in motor-related cortical networks ^1-4^. Systematic reviews suggest overall benefits of MT for post-stroke upper-limb recovery, but implementation heterogeneity and control-condition credibility remain important challenges for trial interpretation and reproducibility ^5-7^. However, MT is not a single task; clinical and experimental implementations vary substantially in movement content, rhythmicity, attentional demands and sensory context. These task features may produce different patterns of task-evoked cortical activation, which in turn may influence mechanistic interpretation and downstream trial design.

Clinically, mirror therapy (MT)—a rehabilitation approach that typically involves mirror visual feedback (MVF)—has been explored as a low-cost, scalable adjunct to conventional rehabilitation for upper-limb recovery after stroke, including early controlled trials and subsequent randomised studies reporting improvements in upper-limb motor outcomes in selected patient groups^8-10^ . Proposed mechanisms include engagement of action observation and motor imagery networks, modulation of sensorimotor excitability via congruent visual feedback, and recalibration of sensorimotor representations when visual input conflicts with impaired proprioceptive/motor output ^3 4^ . Neurophysiological studies in stroke further suggest that MVF-based training can modulate motor-network activity, but reported effects vary with task demands, laterality, lesion characteristics and the quality/credibility of the induced mirror illusion ^11^.

Functional near-infrared spectroscopy (fNIRS) provides a non-invasive and practical approach for quantifying task-evoked haemodynamic responses in superficial cortical regions, including premotor and sensorimotor cortices ^12-16^. A within-participant mechanistic protocol can therefore be used to compare candidate MT task paradigms under standardised conditions, enabling reproducible estimation of neural signatures and subjective experience associated with each task paradigm ^17 18^.

This is an acute, mechanistic, within-participant study designed to characterise neural and experiential differences across task paradigms; it is not intended to evaluate clinical efficacy or long-term outcomes. The primary objective is to compare ROI-level HbO activation (GLM-derived β estimates) across UMT1, UMT2 and UMT3 in healthy adults under a prespecified task-based fNIRS acquisition and analysis framework. Secondary objectives are to compare (1) ΔHbR and other exploratory fNIRS-derived metrics (where prespecified) and (2) post-condition subjective experience ratings across UMT1–UMT3.

## Methods and analysis

### Study design

This will be a single-visit, within-participant, randomised crossover study. Each participant will complete all three conditions (UMT1/UMT2/UMT3) in a Latin-square counterbalanced order to minimise systematic order effects. Reporting will consider SPIRIT guidance with adaptations appropriate for mechanistic crossover experiments^19 20^ and relevant CONSORT guidance for crossover and nonpharmacologic trials ^21-23^.

### Trial status

At the time of manuscript submission, recruitment and data collection have not yet started.

### Setting

The study will be conducted at Wuhan Wuchang Hospital (Rehabilitation Medicine Department), Wuhan, China, in a single standardised rehabilitation training room that also serves as the fNIRS data acquisition room.

### Participants

#### Recruitment

Participants will be recruited via WeChat-based electronic posters and referral through personal networks (word of mouth). Eligibility will be confirmed through screening (telephone prescreening and on-site assessment, as applicable). Written informed consent will be obtained prior to any study procedure.

#### Inclusion criteria

Aged 18–35 years, male or female.

Right-handed (Edinburgh Handedness Inventory score ≥60%).

No known history of neurological disorders (e.g., stroke, epilepsy, Parkinson’s disease).

No history of major chronic diseases affecting cerebral haemodynamics (e.g., uncontrolled hypertension, diabetes, severe dyslipidaemia).

Normal or corrected-to-normal vision and hearing; no colour blindness/colour weakness affecting visual tasks.

Scalp intact without lesions; able to safely wear the fNIRS cap/optodes.

Able to understand study procedures and provide written informed consent.

#### Exclusion criteria

Long-term use of centrally acting medications.

Participation in fNIRS/fMRI/EEG or mirror-therapy related experiments within the previous 3 months.

Head circumference <54 cm or >58 cm.

Pregnancy or lactation.

Alcohol consumption within 24 hours, or caffeine/energy drinks within 6 hours prior to the session.

Any condition preventing completion of the full session or safe task execution, as judged by the investigator.

### Sample size

A total sample size of 24 healthy participants is planned, allowing for approximately 20 analysable participants after accounting for potential data loss. The sample size is based on G*Power calculations for repeated-measures designs (power 0.80, two-sided α=0.05, medium effect size Cohen’s f=0.25, and within-participant correlation ρ=0.5), using a repeated-measures ANOVA framework as an approximation for the planned within-participant comparisons ^24^.

### Randomisation and blinding

The order of UMT1/UMT2/UMT3 will be counterbalanced across participants using a 3×3 Latin-square scheme. The randomisation list will be generated in advance by a team member not involved in outcome analysis and stored as a locked allocation table. The experimenter will implement sequence assignment sequentially by enrolment order; the assigned sequence ID will be recorded in the session log.

This is an open-label mechanistic protocol. Participants will be informed they will complete three different task conditions but will not be informed of any hypotheses regarding which task is expected to induce greater cortical activation. Outcome assessors (for any behavioural scoring beyond automated fNIRS processing) and data analysts will work on de-identified datasets with condition labels encoded.

In operational logs, conditions may be recorded as Run1–Run3; these correspond to the three counterbalanced conditions (UMT1–UMT3) and will be mapped according to the assigned Latin-square sequence prior to analysis.

### Interventions

All conditions will be administered in a standardised MT setup (mirror size approximately 35×45 cm; mirror placed vertically along the midline of the body on the table surface; the left upper limb positioned behind the mirror and fully occluded; the right upper limb positioned in front of the mirror parallel to the mirror surface). The wrist crease will be positioned approximately 3 cm from the mirror edge. All tasks will be performed with the right (dominant) hand to standardise motor execution across conditions.

#### UMT1: task-oriented rhythmic functional movement

Participants will perform a standardised functional task paced by a metronome at 1 Hz: grasping a table-tennis ball with the right hand and placing it into a designated box in synchrony with auditory cues (pure tone ∼1000 Hz, 50 ms duration).

#### UMT2: open-ended free movement

Participants will perform continuous, non-rhythmic, unconstrained right-hand movements (e.g., natural movement and grasping) with the requirement that pauses do not exceed 3 seconds. No rhythmic metronome pacing is provided. To balance auditory input across conditions, continuous white noise at a matched overall loudness to the UMT1/UMT3 metronome cues will be presented throughout the task period without providing rhythmic movement guidance. To enhance standardisation of UMT2, operators will provide prespecified instructions and will record any pauses >3 s or major deviations in the session log; these deviations will be considered in prespecified sensitivity analyses.

#### UMT3: non-functional rhythmic movement

Participants will perform rhythmic fist opening/closing (alternating fist and open hand) paced by an identical metronome cue to UMT1 (1 Hz; same tone parameters).

### Task paradigm

#### Overall structure

Each session will include a 6-minute resting-state baseline fNIRS recording, followed by three task conditions (UMT1/UMT2/UMT3) that will each be performed once. For each condition, participants will complete a 60-second rest/adaptation period and then five task–rest blocks (20 seconds of task followed by 20 seconds of rest), resulting in a total condition duration of 260 seconds (60 seconds rest/adaptation + 5×(20 seconds task + 20 seconds rest)). The three conditions are acquired discontinuously in separate runs (the fNIRS cap remains in place throughout the session). Immediately after each run, fNIRS acquisition will be paused while participants complete a brief subjective experience questionnaire (see Subjective experience assessment).A 2-minute rest interval will be applied between conditions to allow haemodynamic recovery; this interval is intended as a pragmatic washout, and any residual period-related effects will be addressed through Latin-square counterbalancing and inclusion of period as a fixed effect in the primary model.

#### Subjective experience assessment

After each condition, participants will complete a brief subjective experience questionnaire with four dimensions rated on a 7-point Likert scale (1=strongly disagree, 7=strongly agree): illusion, immersion, confusion, and fatigue.

#### fNIRS acquisition

fNIRS data will be acquired using the NirSmart-6000A system (Danyang Huichuang Medical Equipment Co., Ltd., Jiangsu, China) at a sampling rate of 11 Hz with wavelengths of 730 and 850 nm. The mean source–detector distance will be 3.0 cm (range 2.7–3.3 cm). The optode configuration will include 23 source and 15 detectors forming 49 channels. Optodes will be positioned based on the international 10–20 (extended) system to cover bilateral motor-related regions, including SM1/M1 and premotor cortex (PMC), with additional coverage over PFC and SMA as per the fixed montage.

A study-specific montage figure with channel numbering (Supplementary Figure S1) and a prespecified channel-to-ROI mapping table (Table A1) will be provided and finalised before analyses. The prespecified primary ROIs are bilateral SM1/M1 and bilateral PMC. ROI-level estimates will be computed as the mean across valid channels mapped to each ROI; ROI×condition values will be set to missing if <50% of ROI-mapped channels remain valid after QC. All acquisition and preprocessing software versions will be documented.

### Channel-to-ROI mapping and optode montage

#### ROI aggregation rule

For each condition, ROI-level HbO activation will be computed as the **mean** across **valid channels** mapped to that ROI (after QC). If fewer than **50%** of ROI-mapped channels are valid for a given ROI×condition, that ROI×condition value will be set to missing.

#### Operational definition of SM1/M1

For ROI-based analyses, **SM1/M1 is operationalised as the union of M1 and S1 channels** (SM1 = M1 ∪ S1), consistent with the fixed mapping below.

**Table A1.** Prespecified channel-to-ROI mapping for the primary ROIs. SM1/M1 was defined as M1 ∪ S1.

| ROI | Hemisphere | Approx. 10–20 anchor | Channels (finalised list) |
| --- | --- | --- | --- |
| SM1/M1 | Left | C3/CP3 | CH36, CH39, CH40 |
| SM1/M1 | Right | C4/CP4 | CH27, CH28, CH30 |
| PMC | Left | FC3/F3 | CH34, CH35, CH37, CH38, CH41, CH49 |
| PMC | Right | FC4/F4 | CH13, CH14, CH25, CH26, CH29, CH48 |

**Supplementary Figure S1.**
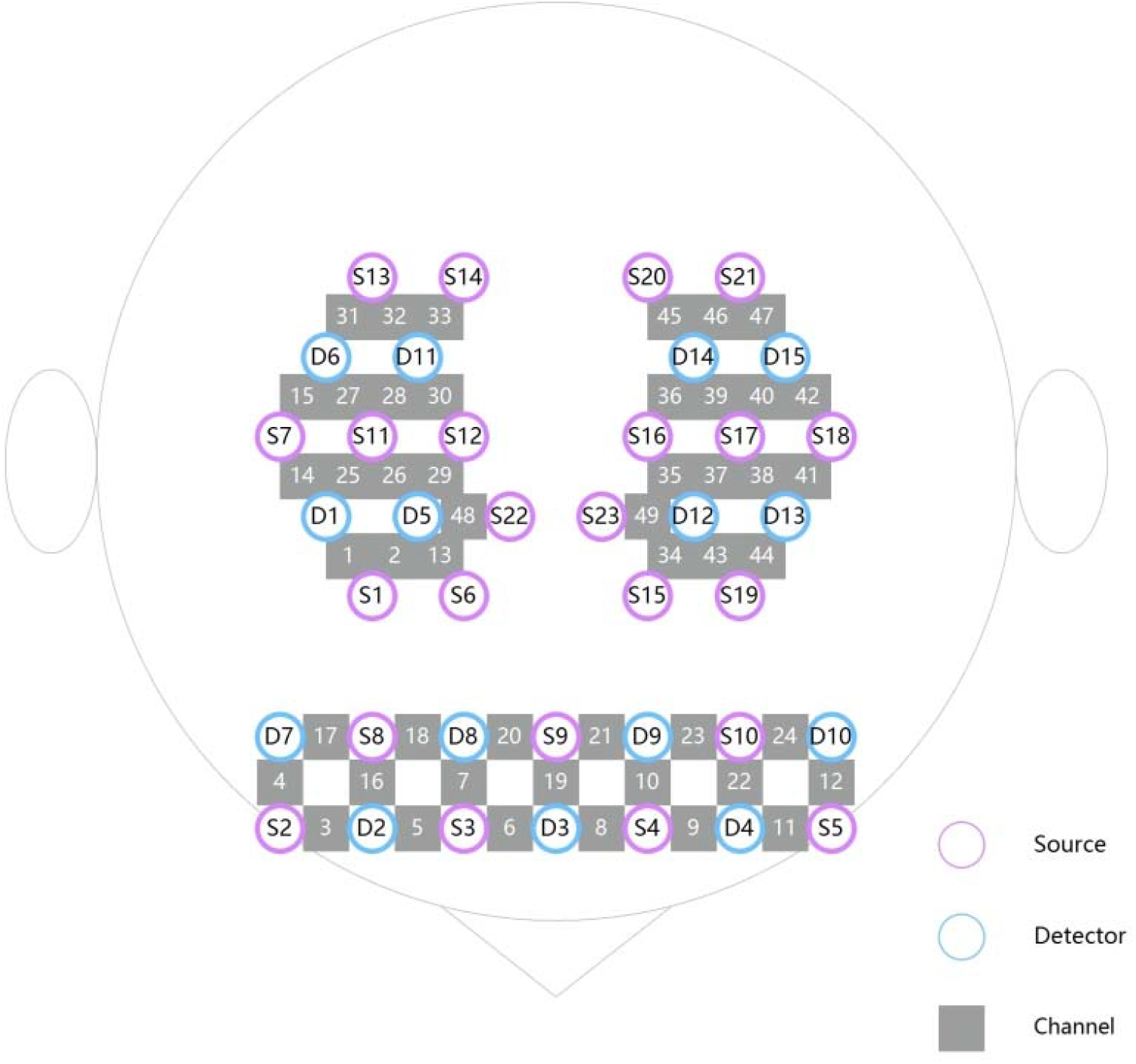
Study-specific fNIRS optode montage with channel numbering. Channel IDs correspond to the prespecified channel-to-ROI mapping (Table A1).

### Quality control

Acquisition-time quality control will be performed by trained operators. Motion, speech and other major artefacts will be time-stamped in the session log ^25-27^. Given known risks of false positives/negatives and physiological confounds in fNIRS, our QC and prespecified preprocessing choices are intended to reduce analytic flexibility and improve interpretability^28-30^ . Online monitoring will flag low quality based on prespecified criteria, including signal quality metrics (SCI < 0.60 and/or CV > 25%) and, where applicable, channel-level criteria (SNR <10 dB and/or motion artefact proportion >30% during a condition). If a channel is flagged as low quality, optodes and hair will be adjusted and QC will be repeated; channels will be marked as bad and excluded from ROI aggregation if still failing QC after troubleshooting. At the condition level, if >20% channels are bad after troubleshooting, that condition will be re-acquired once after adjustment; if still failing, the condition will be marked invalid. Participants will be included in the primary ROI analysis only if at least 2 of the 3 conditions are valid. An ROI×condition datapoint will be set to missing if <50% ROI-mapped channels remain valid after QC.

### Preprocessing and outcome computation

Planned preprocessing includes motion artefact correction ^25-27^, conversion using the modified Beer–Lambert law, and band-pass filtering (0.01–0.20 Hz) applied to HbO/HbR time series^15 31^ . Where short-separation measurements are available, we will consider including short-separation regressors to reduce superficial/systemic contamination in task-based haemodynamic estimates ^32 33^. Task-evoked activation will be estimated using a general linear model (GLM) with a boxcar task function convolved with a canonical haemodynamic response function ^34-36^. All preprocessing and modelling steps will be scripted and version-controlled to support reproducibility, with key analysis code and software versions documented ^29 30^. ROI definitions will be prespecified. ROI-level activation will be summarised as the mean of channel-level estimates within each ROI.

### Outcomes

#### Primary outcome

The primary outcome is ROI-level task-evoked activation in oxygenated haemoglobin (HbO), estimated as GLM-derived β estimates for each condition within the prespecified primary ROIs (bilateral SM1/M1 and bilateral PMC). Primary estimation will be based on these GLM-derived β estimates, which will serve as the main inputs for inference.

#### Secondary and exploratory outcomes

As a complementary descriptive summary, ROI-level HbO will also be summarised as windowed ΔHbO within a prespecified post-onset window of 5–20 s relative to the immediately preceding rest period (descriptive only). Secondary and exploratory outcomes include ROI-level task-evoked changes in deoxygenated haemoglobin (ΔHbR) within the same prespecified ROIs, post-condition subjective experience ratings (illusion, immersion, confusion and fatigue; 1–7 Likert), and exploratory functional connectivity metrics derived from HbO signals (where prespecified). Any exploratory analyses (including connectivity) will be prespecified prior to analysis and clearly labelled as exploratory in reporting ^37^.

#### Statistical analysis plan

All primary inferential analyses will be based on ROI-level GLM-derived HbO β estimates within the prespecified primary ROIs (bilateral SM1/M1 and bilateral PMC). Condition effects will be analysed using a linear mixed-effects model with fixed effects for condition (UMT1/UMT2/UMT3; UMT3 as the reference) and period (1st/2nd/3rd position). Random intercepts will be specified for participant and ROI. Age and sex will be included as covariates.

The primary comparison will test the overall condition effect, followed by prespecified pairwise contrasts with multiplicity control. Planned post hoc pairwise comparisons will use Bonferroni correction (two-sided α=0.05; adjusted α=0.017 for three comparisons). Multiplicity control will be applied to the prespecified family of comparisons (contrast set × ROI) and applied consistently.

Subjective ratings will be analysed using Friedman tests, with Dunn–Bonferroni post hoc tests when appropriate. Pearson correlation will explore associations between PMC HbO β estimates and illusion ratings. Missing ROI×condition values resulting from QC will not be imputed. Statistical analyses will be conducted in R (version 4.4.1).

#### Data management and confidentiality

The study will use de-identified study codes. The re-identification key will be stored separately with restricted access. Data will be recorded on standardised case report forms and managed using the ResMan electronic data capture platform with a full audit trail. After database hard lock, all fields become read-only with a full audit trail of modifications (user, timestamp, old value, new value, reason).

#### Data availability

De-identified participant data will be made available in accordance with ethics approval and institutional policy via a public repository after publication of the primary results; access will require a methodologically sound proposal and a data use agreement prohibiting re-identification.

## Discussion

This protocol is designed as an acute, within-participant mechanistic screening study to compare three prespecified upper-limb mirror-therapy task paradigms (UMT1–UMT3) under tightly standardised acquisition, processing and analytic assumptions. Mirror therapy is widely implemented in clinical practice, yet the “active ingredients” of MT may differ across task implementations that vary in functional goal-directedness, rhythmicity and attentional demand^5-7^. By combining a Latin-square counterbalanced crossover design with task-fNIRS, the present study aims to provide a reproducible framework for characterising how these task features relate to cortical haemodynamic responses and participant-reported experience^15 16^. Importantly, the objective is not to infer clinical efficacy; rather, the protocol is intended to reduce uncertainty when selecting and standardising task paradigms for subsequent translational studies and trials. This protocol is hypothesis-generating and is not designed or powered to evaluate clinical efficacy or long-term treatment effects.

The planned interpretation emphasises convergent mechanistic evidence across physiological and experiential domains. The primary inferential outcome is ROI-level HbO activation quantified as GLM-derived β estimates within prespecified primary ROIs (bilateral SM1/M1 and bilateral PMC), which provides a model-based summary aligned with the block-design timing^34-36^. Complementary windowed ΔHbO summaries and HbR metrics are intended to improve interpretability and to support transparent reporting, but are not positioned as the primary input to hypothesis testing. Alongside fNIRS-derived outcomes, brief post-condition subjective ratings (illusion, immersion, confusion and fatigue) will provide a pragmatic assessment of participant experience that may help contextualise neurophysiological contrasts, inform feasibility and guide refinement of task instructions. Together, these outcomes are expected to support mechanistic hypotheses about how task structure and sensory context may shape engagement of motor-related cortical networks during MT-like training.

Several design features are intended to strengthen internal validity and reproducibility. The within-participant crossover design reduces between-participant variability and increases efficiency for mechanistic comparisons, while the Latin-square counterbalancing and explicit modelling of period effects mitigate systematic order-related confounding. The protocol prespecifies key analytic choices that are frequent sources of flexibility in fNIRS studies, including task timing, primary ROIs, a priori channel-to-ROI mapping, ROI aggregation and missingness rules, and multiplicity control for planned contrasts^28-30^. In addition, the operational separation between execution labels (Run1–Run3) and analytical condition labels (UMT1–UMT3) provides a practical safeguard against documentation errors, because the mapping is determined by the locked randomisation sequence and verified prior to analysis. Finally, the QC and re-acquisition rules were designed to create a closed-loop process (channel → condition → participant) that balances data quality, feasibility and transparency.

This study also has limitations that should be considered when interpreting subsequent results and when planning translation to clinical populations. First, the study is conducted in healthy adults in a single session; neural responses and subjective experience in stroke populations may differ due to lesion location, motor impairment, altered interhemispheric dynamics and differences in attention or fatigue^5 9-11^. Second, fNIRS is restricted to superficial cortical regions and remains sensitive to scalp–optode coupling and motion-related artefacts^15 26 32 33^. Although the protocol incorporates prespecified online monitoring and troubleshooting, residual artefacts and physiological noise may persist and could differentially affect conditions that elicit more movement or require more postural adjustments. Third, despite counterbalancing and inclusion of period as a fixed effect, learning, habituation, fatigue and carryover cannot be eliminated fully in a single-visit crossover design; these influences may be particularly relevant for paradigms that differ in rhythmic pacing or goal-directedness. Fourth, the open-ended nature of UMT2, while intentionally selected to capture a less constrained movement context, may introduce variability in movement kinematics and cognitive strategy across participants; this variability may attenuate or complicate mechanistic contrasts unless adherence and deviations are carefully documented.

The protocol addresses several potential biases and confounders through both design and operational procedures^6 7 22 38^. To reduce expectation-related effects, participants will be informed that they will complete three different conditions without being told any hypothesis about which is “better” or expected to elicit greater activation. Auditory exposure is balanced across conditions through matched cueing/control sounds, aiming to reduce confounding by sensory input. Motion, speech and other major events are time-stamped in session logs and handled through prespecified QC thresholds, bad-channel exclusion, re-acquisition allowances and ROI-level missingness rules. The primary analysis incorporates period effects, and any remaining concerns about order or carryover may be explored through sensitivity analyses. While these strategies cannot remove all sources of bias inherent to behavioural task-fNIRS studies, they support transparent, rule-based handling of common threats to validity.

If this mechanistic screening protocol identifies robust and interpretable differences in ROI-level activation and/or subjective experience across UMT1–UMT3, the findings could inform the selection and standardisation of MT task paradigms for subsequent studies. Next steps may include replication in stroke cohorts, evaluation of test–retest reliability and feasibility in patients with varying impairment levels, and integration of behavioural kinematics and clinical outcomes to assess whether mechanistic signatures during task-fNIRS correspond to meaningful functional changes^5 11 30^. More broadly, the present protocol provides a template for separating intervention “content” (task structure and sensory context) from delivery “format” (mirror setup) and for prespecifying analytic decisions that enhance reproducibility, thereby supporting a clearer mechanistic basis for MT-related trial design.

## Ethics and dissemination

Ethics approval was obtained from the Ethics Committee of Wuchang Hospital Affiliated to Wuhan University of Science and Technology (Approval No.: 2025-112-01; approved on 2025-08-21). This study is expected to be minimal risk. Potential discomforts include scalp pressure from the cap/optodes, mild skin irritation, fatigue from repeated hand movements, and transient dizziness. Adverse events will be recorded and managed according to institutional procedures. Results will be disseminated through peer-reviewed publications and academic conferences.

## Author contributions

Author contributions (CRediT): Conceptualization: Tangzhu Yang, Siyuan Wei, Yuezhu Wang, Dingqun Bai. Methodology: Tangzhu Yang, Siyuan Wei, Yuezhu Wang, Dingqun Bai. Project administration: Tangzhu Yang, Dingqun Bai. Supervision: Tangzhu Yang, Dingqun Bai. Formal analysis: Tangzhu Yang, Siyuan Wei, Yuezhu Wang. Data curation: Tangzhu Yang, Siyuan Wei. Writing – original draft: Tangzhu Yang, Siyuan Wei. Writing – review & editing: Tangzhu Yang, Siyuan Wei, Dingqun Bai. Visualization: Tangzhu Yang, Siyuan Wei. Funding acquisition: Tangzhu Yang, Dingqun Bai.

## Patient and public involvement

Patients and the public were not involved in the design of this mechanistic screening protocol in healthy participants.

## Consent to participate

Written informed consent will be obtained from all participants before any study procedure.

## Patient consent for publication

Not applicable.

## References

1. Ramachandran VS. Phantom limbs, neglect syndromes, repressed memories, and Freudian psychology. Int Rev Neurobiol 1994;37:291–333; discussion 69-72. doi: 10.1016/s0074-7742(08)60254-8

2. Ramachandran VS, Rogers-Ramachandran D. Synaesthesia in phantom limbs induced with mirrors. Proceedings of the Royal Society of London Series B: Biological Sciences 1997;263(1369):377–377. doi: 10.1098/rspb.1996.0058

3. Rizzolatti G, Craighero L. The mirror-neuron system. Annu Rev Neurosci 2004;27:169–92. doi: 10.1146/annurev.neuro.27.070203.144230

4. Caspers S, Zilles K, Laird AR, et al. ALE meta-analysis of action observation and imitation in the human brain. Neuroimage 2010;50(3):1148–1148. doi: 10.1016/j.neuroimage.2009.12.112 [published Online First: 20100104]

5. Thieme H, Morkisch N, Mehrholz J, et al. Mirror therapy for improving motor function after stroke. Cochrane Database Syst Rev 2018;7(7):Cd008449. doi: 10.1002/14651858.CD008449.pub3 [published Online First: 2018/07/12]

6. Hoffmann TC, Glasziou PP, Boutron I, et al. Better reporting of interventions: template for intervention description and replication (TIDieR) checklist and guide. BMJ 2014;348:g1687. doi: 10.1136/bmj.g1687 [published Online First: 20140307]

7. Boutron I, Altman DG, Moher D, et al. CONSORT Statement for Randomized Trials of Nonpharmacologic Treatments: A 2017 Update and a CONSORT Extension for Nonpharmacologic Trial Abstracts. Ann Intern Med 2017;167(1):40–40. doi: 10.7326/m17-0046 [published Online First: 20170620]

8. Altschuler EL, Wisdom SB, Stone L, et al. Rehabilitation of hemiparesis after stroke with a mirror. Lancet 1999;353(9169):2035–2035. doi: 10.1016/s0140-6736(99)00920-4

9. Yavuzer G, Selles R, Sezer N, et al. Mirror therapy improves hand function in subacute stroke: a randomized controlled trial. Arch Phys Med Rehabil 2008;89(3):393–393. doi: 10.1016/j.apmr.2007.08.162

10. Dohle C, Pullen J, Nakaten A, et al. Mirror therapy promotes recovery from severe hemiparesis: a randomized controlled trial. Neurorehabil Neural Repair 2009;23(3):209–17. doi: 10.1177/1545968308324786 [published Online First: 20081212]

11. Rossiter HE, Borrelli MR, Borchert RJ, et al. Cortical mechanisms of mirror therapy after stroke. Neurorehabilitation and Neural Repair 2015;29(5):444–444. doi: 10.1177/1545968314554622

12. Jobsis FF. Noninvasive, infrared monitoring of cerebral and myocardial oxygen sufficiency and circulatory parameters. Science 1977;198(4323):1264–1264. doi: 10.1126/science.929199

13. Ferrari M, Quaresima V. A brief review on the history of human functional near-infrared spectroscopy (fNIRS) development and fields of application. Neuroimage 2012;63(2):921–921. doi: 10.1016/j.neuroimage.2012.03.049 [published Online First: 2012/04/19]

14. Franceschini MA, Fantini S, Thompson JH, et al. Hemodynamic evoked response of the sensorimotor cortex measured noninvasively with near-infrared optical imaging. Psychophysiology 2003;40(4):548–548. doi: 10.1111/1469-8986.00057

15. Scholkmann F, Kleiser S, Metz AJ, et al. A review on continuous wave functional near-infrared spectroscopy and imaging instrumentation and methodology. Neuroimage 2014;85 Pt 1:6–27. doi: 10.1016/j.neuroimage.2013.05.004 [published Online First: 20130516]

16. Pinti P, Tachtsidis I, Hamilton A, et al. The present and future use of functional near-infrared spectroscopy (fNIRS) for cognitive neuroscience. Ann N Y Acad Sci 2020;1464(1):5–29. doi: 10.1111/nyas.13948 [published Online First: 2018/08/08]

17. Qiu Y, Zheng Y, Liu Y, et al. Synergistic Immediate Cortical Activation on Mirror Visual Feedback Combined With a Soft Robotic Bilateral Hand Rehabilitation System: A Functional Near Infrared Spectroscopy Study. Front Neurosci 2022;16:807045. doi: 10.3389/fnins.2022.807045 [published Online First: 20220204]

18. Wei Y, Wu L, Huang F, et al. Effects of robot assisted mirror therapy on motor function and cortical activation in patients with right hemisphere damage. Sci Rep 2025;15(1):33490. doi: 10.1038/s41598-025-16686-y [published Online First: 20250929]

19. Chan AW, Karam G, Pymento J, et al. Reporting summary results in clinical trial registries: updated guidance from WHO. Lancet Glob Health 2025;13(4):e759–e68. doi: 10.1016/S2214-109X(24)00514-X

20. Hrobjartsson A, Boutron I, Hopewell S, et al. SPIRIT 2025 explanation and elaboration: updated guideline for protocols of randomised trials. BMJ 2025;389:e081660. doi: 10.1136/bmj-2024-081660 [published Online First: 20250428]

21. Boutron I, Moher D, Altman DG, et al. Extending the CONSORT statement to randomized trials of nonpharmacologic treatment: explanation and elaboration. Ann Intern Med 2008;148(4):295–295. doi: 10.7326/0003-4819-148-4-200802190-00008

22. Schulz KF, Altman DG, Moher D. CONSORT 2010 statement: updated guidelines for reporting parallel group randomised trials. Bmj 2010;340:c332. doi: 10.1136/bmj.c332 [published Online First: 20100323]

23. McKenzie JE, Taljaard M, Hemming K, et al. Reporting of cluster randomised crossover trials: extension of the CONSORT 2010 statement with explanation and elaboration. BMJ 2025;388:e080472. doi: 10.1136/bmj-2024-080472 [published Online First: 20250106]

24. Eldridge SM, Chan CL, Campbell MJ, et al. CONSORT 2010 statement: extension to randomised pilot and feasibility trials. Bmj 2016;355:i5239. doi: 10.1136/bmj.i5239 [published Online First: 20161024]

25. Scholkmann F, Spichtig S, Muehlemann T, et al. How to detect and reduce movement artifacts in near-infrared imaging using moving standard deviation and spline interpolation. Physiological Measurement 2010;31(5):649–649. doi: 10.1088/0967-3334/31/5/004

26. Brigadoi S, Ceccherini L, Cutini S, et al. Motion artifacts in functional near-infrared spectroscopy: a comparison of motion correction techniques applied to real cognitive data. Neuroimage 2014;85 Pt 1(0 1):181–91. doi: 10.1016/j.neuroimage.2013.04.082 [published Online First: 20130429]

27. Piper SK, Krueger A, Koch SP, et al. A wearable multi-channel fNIRS system for brain imaging in freely moving subjects. Neuroimage 2014;85 Pt 1(0 1):64–71. doi: 10.1016/j.neuroimage.2013.06.062 [published Online First: 2013/07/03]

28. Tachtsidis I, Scholkmann F. False positives and false negatives in functional near-infrared spectroscopy: issues, challenges, and the way forward. Neurophotonics 2016;3(3):031405. doi: 10.1117/1.NPh.3.3.031405 [published Online First: 20160309]

29. Poldrack RA, Baker CI, Durnez J, et al. Scanning the horizon: towards transparent and reproducible neuroimaging research. Nat Rev Neurosci 2017;18(2):115–115. doi: 10.1038/nrn.2016.167 [published Online First: 20170105]

30. Yucel MA, Luhmann AV, Scholkmann F, et al. Best practices for fNIRS publications. Neurophotonics 2021;8(1):012101. doi: 10.1117/1.NPh.8.1.012101 [published Online First: 20210107]

31. Almajidy RK, Mankodiya K, Abtahi M, et al. A Newcomer’s Guide to Functional Near Infrared Spectroscopy Experiments. IEEE Rev Biomed Eng 2020;13:292–308. doi: 10.1109/rbme.2019.2944351 [published Online First: 2019/10/22]

32. Saager RB, Berger AJ. Direct characterization and removal of interfering absorption trends in two-layer turbid media. J Opt Soc Am A Opt Image Sci Vis 2005;22(9):1874–1874. doi: 10.1364/josaa.22.001874

33. Gagnon L, Cooper RJ, Yucel MA, et al. Short separation channel location impacts the performance of short channel regression in NIRS. Neuroimage 2012;59(3):2518–2518. doi: 10.1016/j.neuroimage.2011.08.095 [published Online First: 20110908]

34. Huppert TJ, Diamond SG, Franceschini MA, et al. HomER: a review of time-series analysis methods for near-infrared spectroscopy of the brain. Appl Opt 2009;48(10):D280–98. doi: 10.1364/ao.48.00d280

35. Hou X, Zhang Z, Zhao C, et al. NIRS-KIT: a MATLAB toolbox for both resting-state and task fNIRS data analysis. Neurophotonics 2021;8(1):010802. doi: 10.1117/1.NPh.8.1.010802 [published Online First: 20210125]

36. Santosa H, Zhai X, Fishburn F, et al. The NIRS Brain AnalyzIR Toolbox. Algorithms 2018;11(5) doi: 10.3390/a11050073 [published Online First: 20180516]

37. Garrison KA, Scheinost D, Finn ES, et al. The (in)stability of functional brain network measures across thresholds. Neuroimage 2015;118:651–61. doi: 10.1016/j.neuroimage.2015.05.046 [published Online First: 20150527]

38. Chan AW, Boutron I, Hopewell S, et al. SPIRIT 2025 statement: updated guideline for protocols of randomised trials. Bmj 2025;389:e081477. doi: 10.1136/bmj-2024-081477 [published Online First: 20250428]

