## Supplementary materials for "A single-session randomised crossover fNIRS study comparing three upper-limb mirror therapy task paradigms in healthy adults: a study protocol"

Supplementary materials (Appendices)

- Appendix A: Channel-to-ROI mapping and optode montage (frozen export).
- Appendix B: TIDieR-lite table for task conditions (UMT1–UMT3).
- Appendix C: Preprocessing parameter freeze table.
- Appendix D: Edinburgh Handedness Inventory (EHI) details.
- Appendix E: Session log (minimum required fields).
- SPIRIT checklist.

**Appendix A: Channel-to-ROI mapping and optode montage**

The optode montage will be positioned according to the international 10–20 (extended) system. Channels will be assigned to ROIs a priori and **kept fixed across all analyses**.

**ROI aggregation rule:** For each condition, ROI-level HbO activation will be computed as the **mean** across **valid channels** mapped to that ROI (after QC). If fewer than **50%** of ROI-mapped channels are valid for a given ROI×condition, that ROI×condition value will be set to missing.

**Mapping source and freezing:** The channel-to-ROI mapping is **pre-specified** and will be **frozen prior to the first participant**. The mapping will be generated from the study-specific NirSmart/NirSpark montage using the built-in channel labeling and ROI/probabilistic registration outputs (with cross-checks against 10–20 anchors). The frozen mapping will be exported (channel list + montage figure), versioned, and stored in the analysis log.

**ROI assignment rule:** ROI assignment will be based on software-generated probabilistic registration outputs (e.g., Brodmann/AAL/LPBA40 labels) anchored to 10–20 landmarks, using the default probabilistic-label rule in NirSpark/NirSpace with manual cross-checks against 10–20 anchors.

**Frozen mapping record:** The exported channel list and montage figure (including software version and export date) will be archived in the analysis log as the frozen mapping record.

**Operational definition of SM1/M1:** For ROI-based analyses, **SM1/M1 is operationalised as the union of M1 and S1 channels** (i.e., SM1 = M1 ∪ S1), consistent with the sensorimotor strip coverage of task-based fNIRS and prespecified here to avoid post hoc ROI switching.

*The final channel-to-ROI mapping table and optode montage figure are provided in the Methods section (Channel-to-ROI mapping and optode montage) and are treated as the frozen record for this protocol.*

**Appendix B: TIDieR-lite table for task conditions (UMT1–UMT3)**

This table standardises the implementation details across the three task conditions.

| Condition | Task definition | Posture & setup | Cueing / pacing | Executor | Adherence / fidelity recording |
|---|---|---|---|---|---|
| UMT1: task-oriented rhythmic functional movement | Grasp a table-tennis ball with the right hand and place it into a designated box in synchrony with cues. | Standardised unilateral mirror setup; left hand occluded and still; right (dominant) hand active. | Metronome 1 Hz (pure tone ~1000 Hz; 50 ms); movement paced by cue. | Trained experimenter/therapist (same executor when feasible). | Session checklist: posture/hand placement; cue started; block timing adhered; major deviations time-stamped (looking away, stopping, incorrect movement). |
| UMT2: open-ended free movement | Continuous white noise at matched overall loudness to UMT1/UMT3 cues, presented throughout the task period without instructing rhythmic movement | Identical to UMT1. | Continuous white noise at matched overall loudness to UMT1/UMT3 cues, presented throughout the task period | Same executor as UMT1. | Operators record pauses >3 s or major deviations in the session log; deviations flagged for prespecified sensitivity analyses. |

| Condition | Task definition | Posture & setup | Cueing / pacing | Executor | Adherence / fidelity recording |
|---|---|---|---|---|---|
| | | | without instructing rhythmic movement. | | |
| UMT3: non-functional rhythmic movement | Rhythmic fist opening/closing (alternating fist and open hand). | Identical to UMT1. | Metronome 1 Hz (same cue parameters as UMT1). | Same executor as UMT1. | Session checklist + time-stamped deviations; confirm cue/timing adherence. |

**Appendix C: Preprocessing parameter freeze table**

The following preprocessing/QC parameters will be **frozen before the first participant** and recorded in the analysis log (NirSpark project export + analysis notebook). Any changes after freezing will require a protocol amendment and version update.

| Module | Item | Primary setting (frozen value) | Freeze time point | Recorded location |
|---|---|---|---|---|
| Motion correction | Detection/correction method | Moving SD-based detection + spline interpolation (primary) | Before first participant | Analysis log + NirSpark export |
| Filtering | Band-pass filter | 0.01–0.20 Hz (primary) | Before first participant | Analysis log + NirSpark export |
| GLM (primary outcome) | HRF | Canonical HRF (software default; version documented) | Before first participant | Analysis log + NirSpark export |
| GLM (primary outcome) | Regressors / covariates | Condition-specific task regressor(s); additional | Before first participant | Analysis log |

| Module | Item | Primary setting (frozen value) | Freeze time point | Recorded location |
|---|---|---|---|---|
| | | covariates (if any) will be prespecified and listed in the analysis log before freezing | | |
| Quality control | SCI threshold | SCI ≥ 0.60 | Before first participant | Protocol (QC section) + analysis log |
| Quality control | CV threshold | CV ≤ 25% | Before first participant | Protocol (QC section) + analysis log |
| Outcome computation | ΔHbO window / baseline (descriptive summary) | 5–20 s post-onset; baseline = immediately preceding rest within each block | Before first participant | Protocol (Outcomes section) + analysis log |

**Appendix D: Edinburgh Handedness Inventory (EHI) details**

Participants' handedness will be assessed using the Edinburgh Handedness Inventory (EHI). The EHI includes 10 common activities; participants indicate their preferred hand for each activity.

EHI items (10 everyday hand-use activities)

- Writing
- Drawing
- Throwing
- Scissors
- Toothbrush
- Knife (without fork)
- Spoon
- Broom (upper hand)
- Striking a match
- Opening a box lid (holding the lid)

Scoring and Laterality Quotient (LQ)

For each item, record whether the participant uses the **Right (R)** hand or **Left (L)** hand (optional: allow “both” only if used consistently; if “both” is allowed, it should be handled according to a prespecified rule).

Compute the Laterality Quotient (LQ) as:

$LQ = ((R - L) / (R + L)) \times 100$.

Handedness threshold for eligibility

Participants will be considered right-handed and eligible if **Edinburgh Handedness Inventory score ≥60%**.

**Appendix E: Session log (minimum required fields)**

This appendix defines the **minimum required** session-log fields to ensure standardised documentation of protocol deviations, interruptions, and data-quality issues across participants and conditions. A printable session-log form may be used; the items below specify the minimum content that must be captured.

Minimum required fields (per participant/session)

- Administrative
  - Study code
  - Session date
  - Start time / end time (or total duration)
  - Site/room (if applicable)
  - Operator/experimenter ID (initials or name)
- Randomisation / condition coding (traceability safeguard)
  - Latin-square **sequence ID**
  - Actual execution order recorded as **Run1–Run3**
  - For each Run, the mapped analytical condition label **UMT1/UMT2/UMT3**
- Protocol deviations / interruptions (time-stamped)
  - Any deviation from standardised instructions (e.g., incorrect posture, looking away, non-compliance)
  - Interruptions (pause/stop), with reason and duration
  - Participant-reported discomfort or request to stop
- fNIRS setup / QC and re-acquisition (time-stamped)
  - Notable optode/cap adjustments (what/when/which side if applicable)
  - QC failures triggering troubleshooting (e.g., low SCI/high CV or other prespecified online QC flags), and corrective actions taken

- Re-acquisition events (which Run/condition, trigger reason, whether it resolved the issue)

- Motion/physiological confounds (time-stamped)
  - Coughing, speaking, visible head/trunk movement
  - Any event judged to compromise block structure or data validity (brief description)
- File/traceability notes
  - Any file-naming exceptions or missing exports (raw/events/QC summaries) and how they were resolved
  - Software/hardware version notes if changed from standard setup

Recording rules

- All major events should be **time-stamped** and should match corresponding event markers where applicable.
- Use the session log to document **what happened**, **when it happened**, and **whether it may affect** a specific Run/block; avoid interpretation beyond factual notes.
